# Characterization of long-term patient-reported symptoms of COVID-19: an analysis of social media data

**DOI:** 10.1101/2021.07.13.21260449

**Authors:** Juan M. Banda, Nicola Adderley, Waheed-Ul-Rahman Ahmed, Heba AlGhoul, Osaid Alser, Muath Alser, Carlos Areia, Mikail Cogenur, Krisitina Fišter, Saurabh Gombar, Vojtech Huser, Jitendra Jonnagaddala, Lana YH Lai, Angela Leis, Lourdes Mateu, Miguel Angel Mayer, Evan Minty, Daniel Morales, Karthik Natarajan, Roger Paredes, Vyjeyanthi S. Periyakoil, Albert Prats-Uribe, Elsie G. Ross, Gurdas Singh, Vignesh Subbian, Arani Vivekanantham, Daniel Prieto-Alhambra

## Abstract

As the SARS-CoV-2 virus (COVID-19) continues to affect people across the globe, there is limited understanding of the long term implications for infected patients^1–3^. While some of these patients have documented follow-ups on clinical records, or participate in longitudinal surveys, these datasets are usually designed by clinicians, and not granular enough to understand the natural history or patient experiences of ‘long COVID’. In order to get a complete picture, there is a need to use patient generated data to track the long-term impact of COVID-19 on recovered patients in real time. There is a growing need to meticulously characterize these patients’ experiences, from infection to months post-infection, and with highly granular patient generated data rather than clinician narratives. In this work, we present a longitudinal characterization of post-COVID-19 symptoms using social media data from Twitter. Using a combination of machine learning, natural language processing techniques, and clinician reviews, we mined 296,154 tweets to characterize the post-acute infection course of the disease, creating detailed timelines of symptoms and conditions, and analyzing their symptomatology during a period of over 150 days.

## Introduction

During the second surge of acute cases of COVID-19 in countries such as the United States and Spain, healthcare systems were overwhelmed with the management of acute cases, giving little attention, at the time, to the impact of infections on survivors and the long-term COVID-19 sequelae. The current wave (Spring 2021) is taking a great toll amongst young people, who are less likely to need hospital or intensive care, but more likely to develop long-term symptoms ^4^. The pathogenesis of post-COVID-19 syndrome (“long-COVID”) is still uncertain, and may simultaneously include sequelae on different tissues caused directly by COVID-19 infection or indirectly by virus-induced immune damage. Initial reports from countries such as Italy^5^ show that many COVID-19 patients were reporting concerning long-term symptoms. These reports mention acute organ injury during primo-infection including acute kidney injury in 20% of patients^5^, myocardial injury in 20%-30%, and acute respiratory failure. Since these initial studies, other clinical reporters like Elisabeth Mahase^6^ and Michael Marshal^7^ started to report similar trends in UK and US patients, respectively. While work on acute patient cohorts was initially limited, more researchers have started to turn their attention to the long-COVID problem^1,2^, which have informed guidelines^8^. Many of these studies have mixed classical post-ICU sequelae with post-COVID-19 symptoms. This is a major limitation of such studies, further complicating the definition of the post-COVID-19 syndrome. Opinion pieces from clinicians^9^ and researchers^10^ have made it clear that anonymous users on the internet, like members of the long-COVID forum^11^, and similar groups in other social media outlets, document their experiences for researchers and medical professionals to analyze. Social media platforms have not been created with health-related purposes in mind^12^, however, people publicly share personal health information every day^13^, thus not confounded by recall bias, representing an important source of health information being spontaneous, unsolicited and up to date. This data from social media can be monitored and analyzed by using natural language processing, providing new ways to better understand users’ health. Twitter is a microblogging social media service platform with more than 330 million active users worldwide^14^. Twitter users post short open available messages about facts, feelings, and opinions, including health conditions ^15^. The COVID-19 pandemic has shown a very unique aspect of involving patient-led initiatives into creating survey-based ‘long-COVID’ studies alongside health care professionals ^16^, or with special interest groups like Survivor Corps ^17^ creating longitudinal survey based studies ^18^ to measure the impacts of post-acute sequelae. Our work is fundamentally different in the sense that we did not directly contact any COVID-19 survivor, patient, or create any survey for people to complete. Instead, we directly identified Twitter messages that use hashtags related to ‘long-COVID’, and ethically^19^ extracted the publicly available timelines for a curated set of users. These timelines narrate the user’s first hand experiences, at their own pace and time, documenting their daily lives alongside their disease sequelae. We leveraged a team of biomedical experts and clinicians to annotate relevant tweets, rather than fully relying on automated machine learning methods for this task. All data points presented in this study are identified by domain experts and have only been standardized by automated methods with a domain expert always involved in review. This work shows that social media data, in particular Twitter combined with the topic of long-COVID, is one additional research data source that could be leveraged alongside clinical and survey data to further characterize and investigate the disease progression.

## Methods Summary

The public Twitter stream is a 1% random sample of all tweets on a given day. We used a combination of tweets from the dataset curated by Banda et.al. ^20^, additional tweets gathered using the Twitter COVID-19 research endpoint ^21^, and our own data collection stream set for long-COVID and related hashtags from 2020-07-04 to 2020-08-10. For these analyses, we included users with three or more tweets in English (within this set), with any of the following hashtags: #LongCovid, #COVIDPERSISTENTE, #longhaulers, #apresJ20, and #CountLongCovid. All retweets were removed at this stage, and never used in our analysis. An important detail to note here is that none of the datasets offer complete timelines of all tweets for any of the selected users. As a starting point we used two clinicians to identify which users are describing post-COVID-19 symptoms. From this cohort we then applied a mixture of domain expert review for tweet annotation, machine learning (ML) for relevance filtering, and natural language processing (NLP) for standardization. Details on these methods are included in the Methods section. Figure 1 outlines our pipeline to generate the data and insights presented in the following sections.

**Figure 1.**
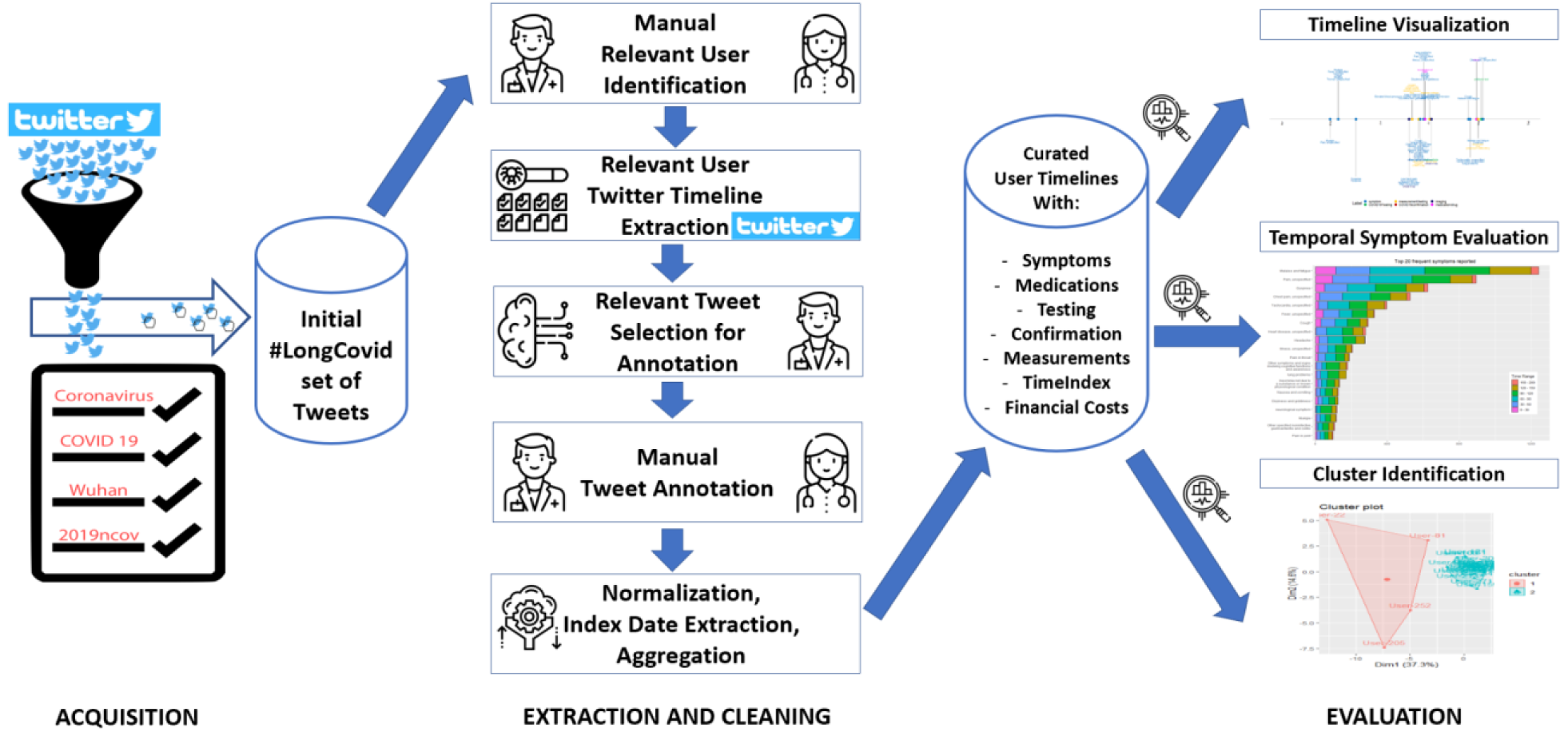
Complete pipeline for the acquisition, extraction, cleaning and evaluation of Twitter user timelines into long-COVID trajectories.

### Tweet gathering, long-hauler identification, and domain-expert annotation

We started with the identification of 8,556 unique users, in a total of 31,481 unique tweets. From which our clinician review identified 312 users after consensus evaluation, detailed in the **Methods: Identification of long haulers in Twitter data**. We then extracted the timelines for 306 of these users, as 6 of them had their account set to private (not publicly visible). From these 306 users, 264,673 tweets were extracted from their Twitter timelines between 2020-01-01 and 2020-08-17. While this number of tweets is unmanageable to have domain-experts curate, we relied on machine learning algorithms to filter only potentially relevant tweets of self-reported COVID-19 symptoms, medications, testing and imaging procedures, additional details can be found in the ***Methods: Extraction and annotation of long-haulers Twitter timelines***. A PRISMA-like diagram is provided as Appendix 1 to detail the record filtering process from start to end.

A total of 12,978 unique tweets were annotated by our domain experts, yielding a total of 31,385 annotations. This step took over 400 domain-expert hours to complete. Additional details on the annotation of the user timeline tweets can be found in the ***Methods: Extraction and annotation of long-haulers Twitter timelines***. Similarly, additional steps were performed to extract the symptom’s index dates to build the patients’ timelines (***Methods: Index date extraction and assignment***) and to normalize the annotations into clinical codes and relevant organ system groups (***Methods: Annotation Normalization***).

### Characterization of self-reported symptoms

It is noteworthy that symptoms are often under-reported in Electronic Health Records (EHR)/Claims datasets as shown in studies across multiple databases from different countries^22,23^, so we believe that our data source contains valuable additional information as it has higher granularity, and it allows for symptoms like brain fog and COVID cheeks, to be identified as there are not ICD10 codes for them. From the cohort of 306 users identified, 286 (93.46%) mentioned at least one symptom in their timeline. The remaining 20 users reported COVID-19 confirmation, testing, medications, or imaging, but no symptoms. From these 286 users, the earliest self-reported symptom date (index date) was 2020-01-06 and the latest was 2020-08-07. All users reported 9.26 distinct symptoms on average, with a minimum of 1 and a maximum 59. In terms of users reporting symptoms over time, Figure 2 shows the distribution of users reporting symptoms over increasing periods of time after COVID-19 infection. To note, we have a high number of users still reporting symptoms between 60 and 120 days from their index date (Figure 2), and 15% of them still reporting symptoms well over five months after reported infection.

**Figure 2.**
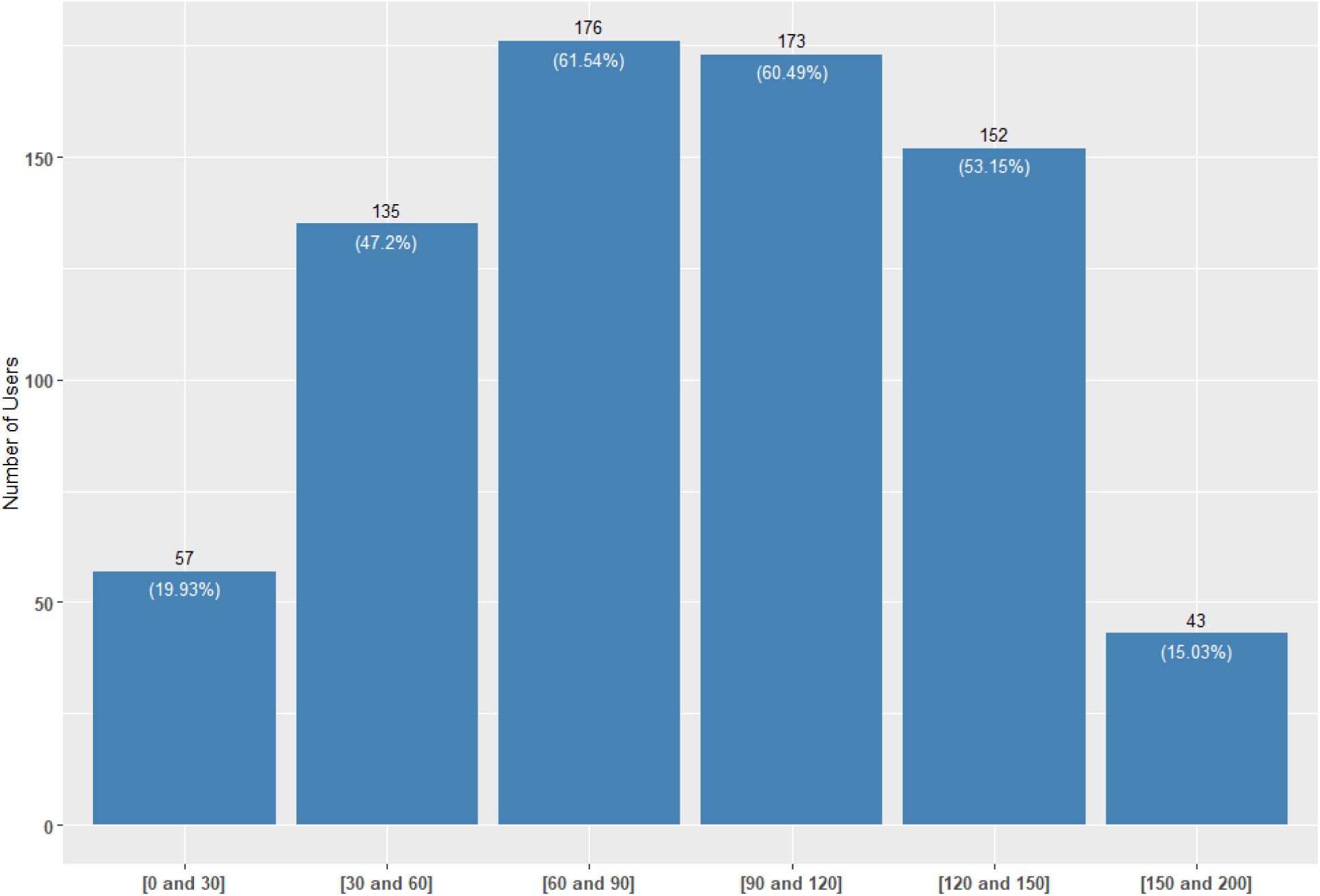
Number of users and percentage of cohort reporting symptoms over time (in days).

Like Carfi and colleagues^24^ in their report of patients hospitalized with COVID-19 in Italy, we also found that the most common long-COVID symptoms are fatigue, dyspnea, joint pain and chest pain, as shown in Figure 3, both in terms of total reports (top panel) and number of users reporting them (bottom panel). Using a mobile application for symptom tracking, Sudre et al.^25^ found that 98% of individuals reported long-term fatigue, and 91% reported intermittent headaches^25^, another finding we are able to identify in our Twitter data. Additionally, we show the temporality of these symptoms in the stacked bars of Figure 3, indicating that most still continue to appear after 120 days.

**Figure 3.**
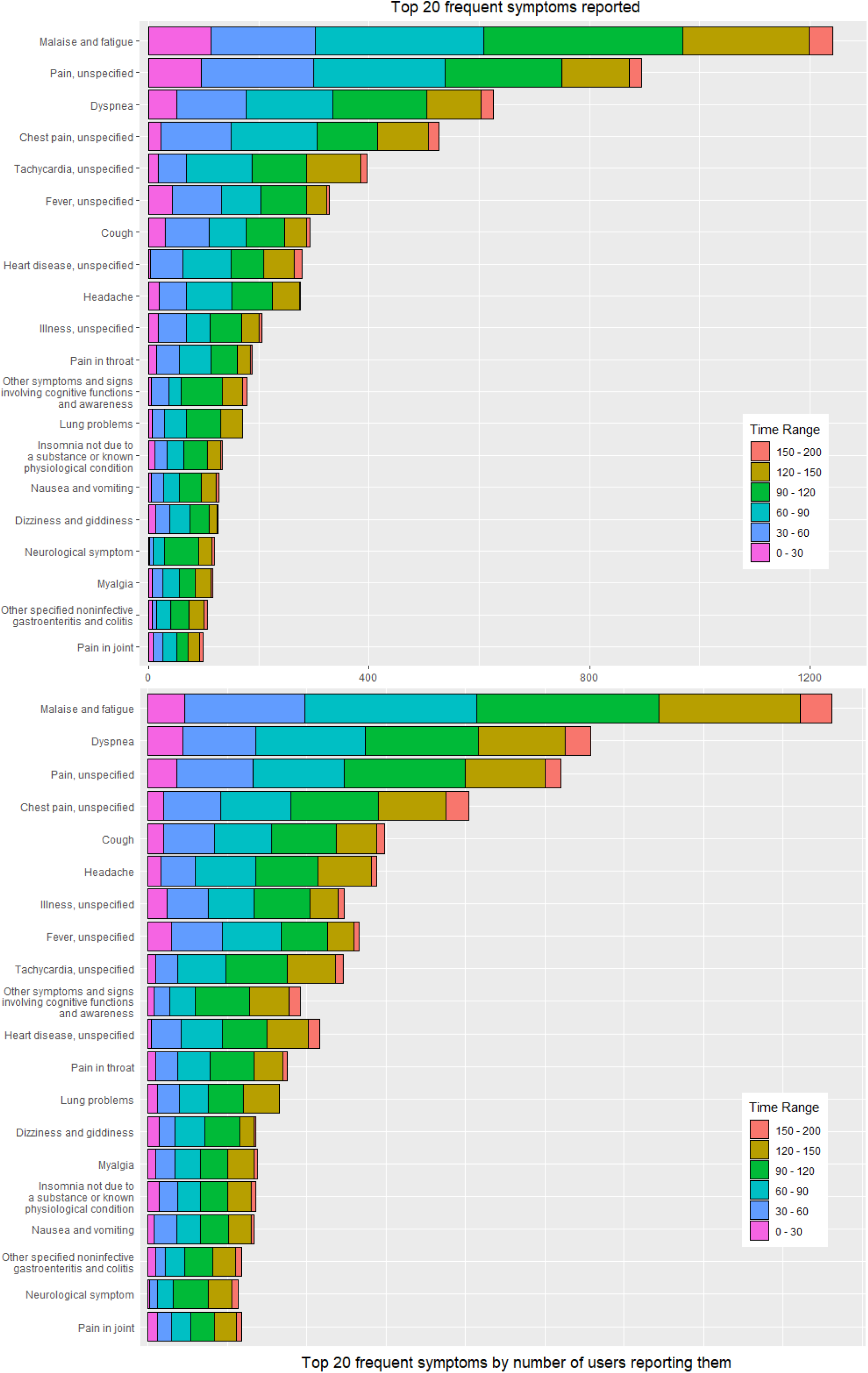
Top 20 symptoms by frequency and user count. Counts are stacked by time ranges, from 0 to 30 days, all the way to 200 days.

In order to show the temporal reporting of any given symptom group, we present Figure 5, which indicates for each symptom group the index days which have reports. Whilst e.g. cardiovascular, respiratory and cutaneous symptoms come later (light yellow, purple and orange symptom groups), others like fatigue, persistent fever/chills and gastrointestinal symptoms (light red) appear early in the patient’s journeys. We also found that fatigue, dyspnea, pain and fever dominated reported symptoms early in an individual’s timeline. However, over time, other symptoms and conditions such as tachycardia, heart disease, and cognitive dysfunction became more commonly reported. This change in the constellation of symptoms and diagnoses reported suggest that COVID-19 may cause irreparable end-organ damage after acute infection, as has been reported in prior studies^26–28^. Also be that we are in the beginning stages of identifying patterns of recovery from infection.

**Figure 5.**
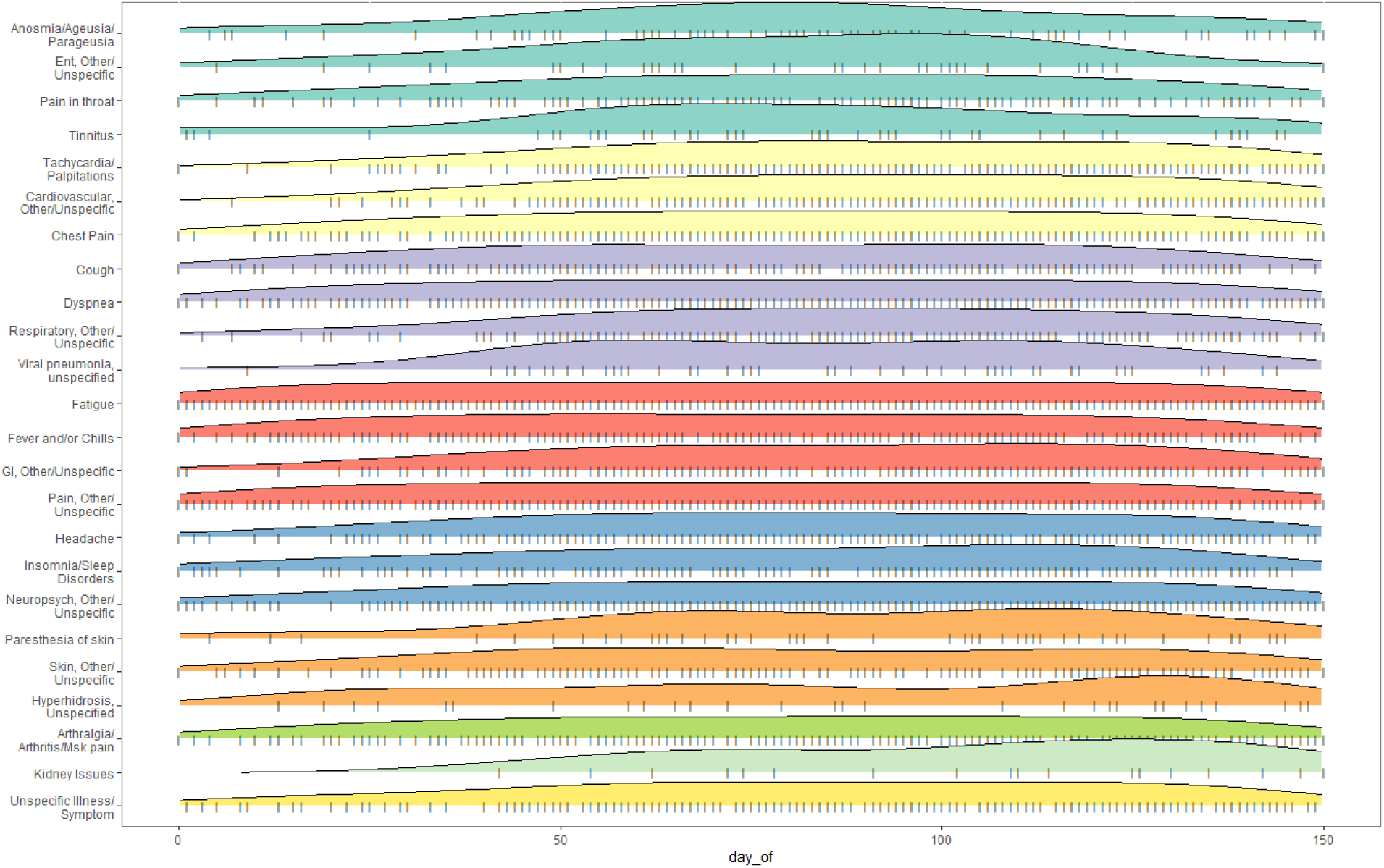
Ridge plot of the temporal distribution of reporting of symptoms. GI is used for Gastro-Intestinal and ENT for Ear, Nose and Throat.

### Measurements, imaging, and medications analysis

While the main focus of this work is characterizing the symptoms of long haulers, we had our clinician team annotate measurements, imaging, and medications to try to uncover any interesting trends. Based on our expert annotations, only 15 out of 306 users (4.9%) mention a clear COVID-19 confirmation. This is not surprising as many users in our cohort were diagnosed at a time when tests were not readily available for those presenting, at the time, mild symptoms. With regards to other tests or measurements being performed, 67 out of 306 users (21.9%) mentioned some sort of measurement. The top five most common measurements or tests and their frequencies are found in Table 1. Blood pressure, oxygen saturation, and heart rate were the top three clinical measurements, in line with the reported cardiovascular and respiratory symptoms above.

**Table 1.**
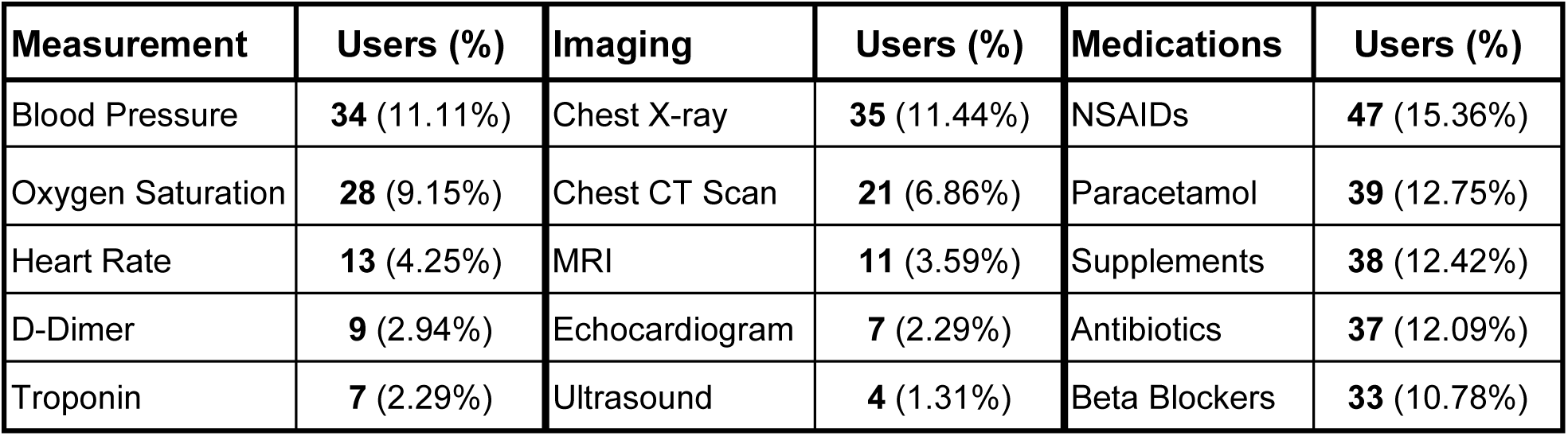
Top five most common measurements, imaging, and medications per user.

Out of 306 users, 36 (11.76%) reported some sort of imaging procedure during their timelines, with respiratory and cardiovascular imaging (chest X-ray/CT, echocardiogram) amongst the most common. In terms of medication intake, 94 out of 306 users (30.72%) mentioned one or more medications during the study time period, including antipyretics and painkillers (NSAIDs, paracetamol), vitamin supplements, antibiotics, and beta blockers.

## Conclusion

Unsurprisingly, the COVID-19 pandemic accelerated a larger trend of increased social media use and engagement, with up to 51% of adults in the United States reporting increased use during the pandemic^29^. As the first major/world-wide pandemic in the times of social media, it has been a great outlet for people to share their experiences during their time being sick, and more importantly after, creating a very large, and detailed set of narratives on their long-COVID journey. We have demonstrated that it is possible to curate Twitter data to perform temporal analyses of self-reported health events. Incorporating traditional methods of manual review and machine learning/data science, we have been able to create a very rich cohort of 306 individuals whose documented experiences provide an extremely valuable insight into the days after their COVID-19 infections. While each narrative is unique in its own way, we have been able to identify symptom patterns within our cohort. We hope that these findings are of help to clinicians and long-COVID clinics in helping their patients through their sequelae, allowing for a proper syndromic definition for long-COVID, and that they are validated in a prospective way in multinational cohorts. Additionally, these can help with improving access to specific healthcare needs like X-rays, etc. It is important to note that the richness and frequency of the user’s symptom descriptions is unparalleled to any other resource for clinical research, as some people post near-real time about their condition. This makes the case to use social media data to capture more longitudinal and frequent data on clinical events that would not be possible using traditional means, and allows us to better understand the progression of diseases and reoccurring or newly occurring symptoms. While the COVID-19 pandemic has brought all these people together to share their journeys online, we also see that other groups of users with chronic illnesses like Myalgic encephalomyelitis/chronic fatigue syndrome (ME/CFS) share similar narratives online, opening future research avenues to explore those cohorts of people and characterize their diseases in a non-invasive way.

### Limitations

Our study has several limitations. First, we rely on patient reported symptoms that are not clinically verified. Nevertheless, this is also a strength, as we aimed to collect near-real time, unbiased patient-reported experiences instead of clinically driven information. We also found that most patients who report long-COVID symptoms never had definitive testing that attributes their illness to COVID-19, which is mostly due to limited testing early in the pandemic. Second, the limitation in using social media data is its extreme unstructured nature, which requires multiple hours of validation by experts. However, we addressed this by building machine learning models that we leveraged to decrease the number of tweets requiring review and adjudication. Lastly, we do not have any information about user comorbidities other than the ones self-disclosed, in contrast to clinical studies. Aside from these limitations, researchers have been using Twitter data in the context of health research for pharmacovigilance^30,31^, toxicovigilance^32^, and more recently to identify cohorts of pregnant women for safety surveillance^33^, using user timelines, showing the potential of this kind of data for health research.

## Methods

### Identification of long haulers in Twitter data

We started with the identification of 8,556 unique users, in a total of 31,481 unique tweets. Once we narrowed down our potential cohort, we grouped all user tweets by user and ordered them sequentially by date. We then presented these groups of tweets per user in an anonymized way (no real user name or tweet ID visible) to two clinicians for determination if the user is talking about a personal long-COVID journey, or if the user is voicing other’s experiences or news/literature mentions of the phenomena. Both clinicians were tasked to make their best assumption based on the set of tweets presented, which varied from 800+ tweets to just two. The average number of tweets presented was 3.7 as we had many users with only a handful of tweets available in our initial dataset. The clinician agreement between potential long haulers was evaluated to be 92%, with the other 8% of disagreements being resolved by a third clinician. A total of 312 people were determined to be good candidates for long haulers based on our manual evaluation. This set of 312 people had an average of 13.4 tweets in our dataset, allowing clinicians to make a confident decision based on multiple data points. Note that there are many more potential long haulers in this dataset we used, however, we wanted to focus on a high-quality and easily identifiable set of users as time and resources for our evaluation were limited.

### Extraction and annotation of long haulers’ Twitter timelines

After having identified the Twitter users for further evaluation, we needed to extract a longitudinal timeline of all of the users’ tweets between the beginning of 2020 (2020-01-01) and the day we started our study (2020-08-17), giving us 229 days to look into the users’ tweeting activity before COVID-19 was declared a pandemic, and plenty of time after the pandemic declaration, to look for persistent symptoms in a span of several months. While the Twitter API only allows free users to extract a very limited set of old tweets, we used two scraping Python packages, GetOldTweets ^34^ and our own scraper part of SMMT^35^ to extract the user timelines. These software packages allowed us to go back all the way to the beginning of the year for each selected user and extract all their tweets over the given period of time. From the 312 users identified, we found that 6 users had their timelines set to private, thus we did not extract their timelines and removed them from the study. From the remaining 306 users we extracted a total of 264,673 tweets. While the average number of tweets per user was ~865, some tweeted with higher frequency than others.

We recruited a set of 24 domain expert annotators for this work, however, annotating 264,673 tweets would have been an unreasonable task as the average annotation rate per hour circled around 90-100 tweets. Also, we had two different domain experts annotate the same set of tweets to calculate inter annotator agreement for quality control. This process would have taken over 5,880 domain expert hours. In order to identify relevant tweets and reduce the annotation time by presenting highly probable relevant tweets to the annotators, we built a machine learning model based on the annotation of an initial set of 150 relevant and 2,453 irrelevant tweets from related earlier work ^36^. We further enhanced this by having our senior clinician reviewer manually annotate an additional set of 3,386 tweets (with 474 relevant and 2,912 irrelevant), giving us a total of 624 relevant and 5,365 irrelevant tweets. Building such models is standard practice ^37^ and we experimented with different classification algorithms and class imbalance scenarios until we trained a support vector machine (SVM) model that performed a 75% accurate classification of a 20% random held-out test set from our manually labeled set. The intricate details of this process were out of the scope of this paper, as we focus on evaluating what was annotated/extracted from the tweets based on the clinician annotations. Since we split our manual review of the extracted timelines into three different rounds, we improved our model in each iteration, achieving 81.5% accuracy for all predicted tweets as relevant with a probability higher than 85%. In each iteration we selected only the tweets with a probability of 70% or higher to be relevant. As an interesting observation, we found that in our initial relevancy annotated set, tweets were at least 110 characters or longer (on average), when compared to the non-relevant tweets which had an average length of 85 characters. All tweets were randomly sorted and fully anonymized (no username or identifier) before they were made available to our group of clinicians for annotation.

Our clinicians were tasked with identifying ten different things in each tweet: COVID-19 Testing mentions, COVID-19 confirmations, mentions of financial costs related to the disease, Time Index mentions of when the user says he got COVID-19, any mentions of a clinical condition, any mentions of Imaging studies performed, any mentions of body measurements or tests, any mentions of medications they are taking, any mentions of disease symptoms, and lastly, if the tweet was not relevant, it would me marked as such. The annotation task was carried out in batches of 700-900 tweets, each one annotated by three separate clinicians. Using the docanno open source text annotation tool ^38^, the annotator was tasked with selecting the section (sequence labeling) of the tweet’s text that best represented any of the given labels. Figure S1 gives an example of the annotation interface with some symptoms and time index annotations.

**Figure S1.**
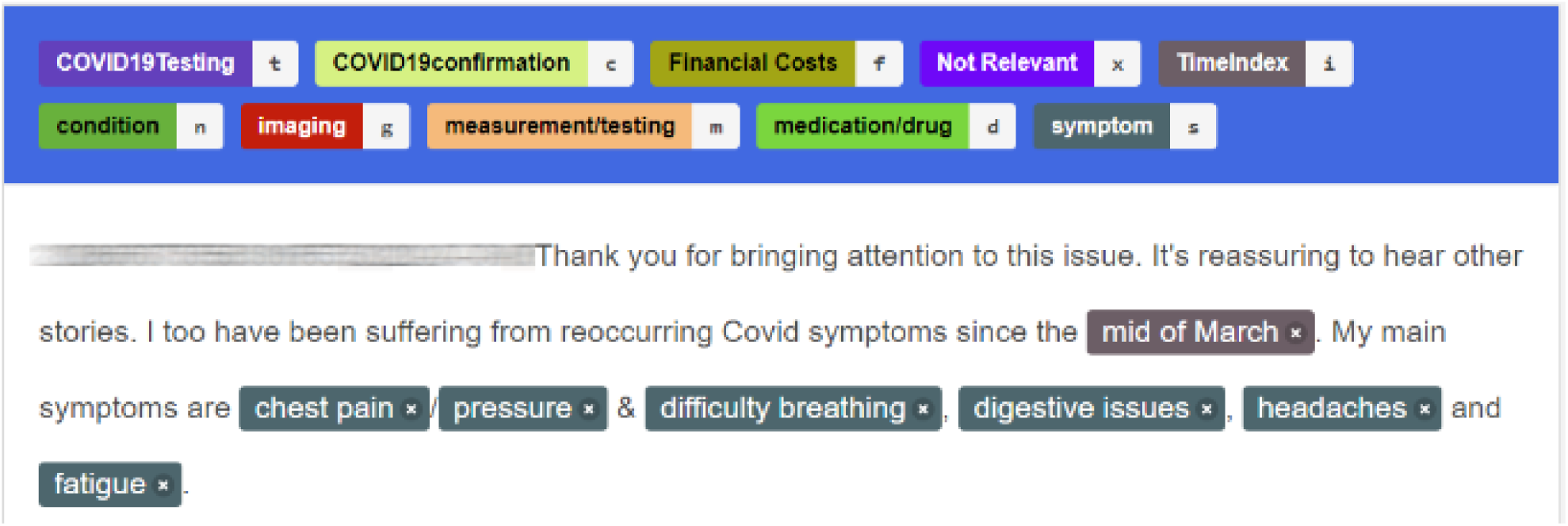
Docanno annotation tool example.

Each tweet was annotated by at least two domain experts. To evaluate the agreement between annotators, we looked at label-level agreement between our clinician reviewers within their allocated sets of tweets. We calculated the pairwise Cohen kappa ^39^ statistic (κ). If both annotators are in complete agreement then κ = 1, in the worst case, if there is no agreement among the annotators (other than what would be expected by chance) then κ ≤ 0 ^40^. Table S1 shows the total number of annotations produced for every single one of our 10 classes, and their average Cohen kappa statistic. Note that these numbers are not unique annotations, but total annotations produced for each category.

**Table S1.** Annotation statistics

| Label | Annotations | Avg. Cohen's kappa - $\kappa$ |
| --- | --- | --- |
| COVID19Testing | 336 | 0.7124 |
| COVID19confirmation | 307 | 0.8143 |
| Financial Costs | 66 | 0.7069 |
| Not Relevant | 8,469 | 0.8671 |
| TimeIndex | 3,381 | 0.8429 |
| Imaging | 227 | 0.8612 |
| Measurement/testing | 1,038 | 0.7935 |
| Medication | 954 | 0.8544 |
| Condition / Symptom | 16,607 | 0.8302 |

A κ of 0.61 and higher is considered substantial agreement and all our labels had a κ value of 0.67 at a minimum. In fact, the κ value for most labels of interest was above 0.70, with some even reaching values above 0.81, which is considered near perfect agreement ^40^.

### Index Date extraction and assignment

In order to perform temporal data analysis for long haulers’ symptoms, we had our clinician annotators label parts of tweets that identified how many days/weeks/months the person had been experiencing symptoms, the day they estimated they got COVID or the actual day they tested positive for COVID, with the *TimeIndex* label. One interesting fact of tweets from long haulers is that many of them keep a very concise track of the number of days they have been experiencing symptoms, or report the actual date of their positive tests. Table S2 shows different variations of the text found by the TimeIndex label. Our calculated index date is highlighted in bold, given the different adjudications scenarios we discuss below.

**Table S2.** Index Date calculation for users, based on their TimeIndex annotations.

| userName | tweetDate | annotationText | Calculated Index Date |
| --- | --- | --- | --- |
| User-103 | 2020-06-01 | day 75 | **2020-03-18** |
| User-103 | 2020-07-17 | 17 weeks in | 2020-03-20 |
| User-105 | 2020-05-09 | 14 days | 2020-04-25 |
| User-105 | 2020-07-05 | after 3 months | 2020-04-06 |
| User-105 | 2020-07-14 | i first had covid on 24th april, | **2020-04-24** |
| User-106 | 2020-05-24 | 60plus-day | 2020-03-25 |
| User-106 | 2020-05-28 | day67 | 2020-03-22 |
| User-106 | 2020-07-01 | 100days | 2020-03-23 |

With over 3,381 TimeIndex labels annotated, we created a custom Python software utility, part of SMMT ^35^ to calculate the potential date of COVID-19 start for each user based on the text from the annotated label. While the software we created gives us an automatically calculated date based on the text, being a specific date, a number of days, or a weeks/months estimate, there will be decisions to adjudicate dates or adjust manually. Our algorithm’s rules for index date assignments are: If the user reports a specific date, use the given date, if the user reports multiple different day estimates, subtract days from the current tweet date and select the one with the highest amount of repetition. If there is an uneven number of reports, pick the average date, lastly, if the user did not report any specific date, number of days, or weeks estimate, use the date of the first tweet with symptoms as the index date. Table S2 shows in the last column the Index date assignment based on our algorithm. Out of our set of 306 long haulers, 282 (~90%) had one or more TimeIndex labels, allowing for a better Index Date estimation.

Once our domain expert-reviewed standardized annotations and the user’s index dates were calculated, we were able to visualize each user and their long hauler journey with plots as shown in Figure S2 A and B.

**Figure S2.**
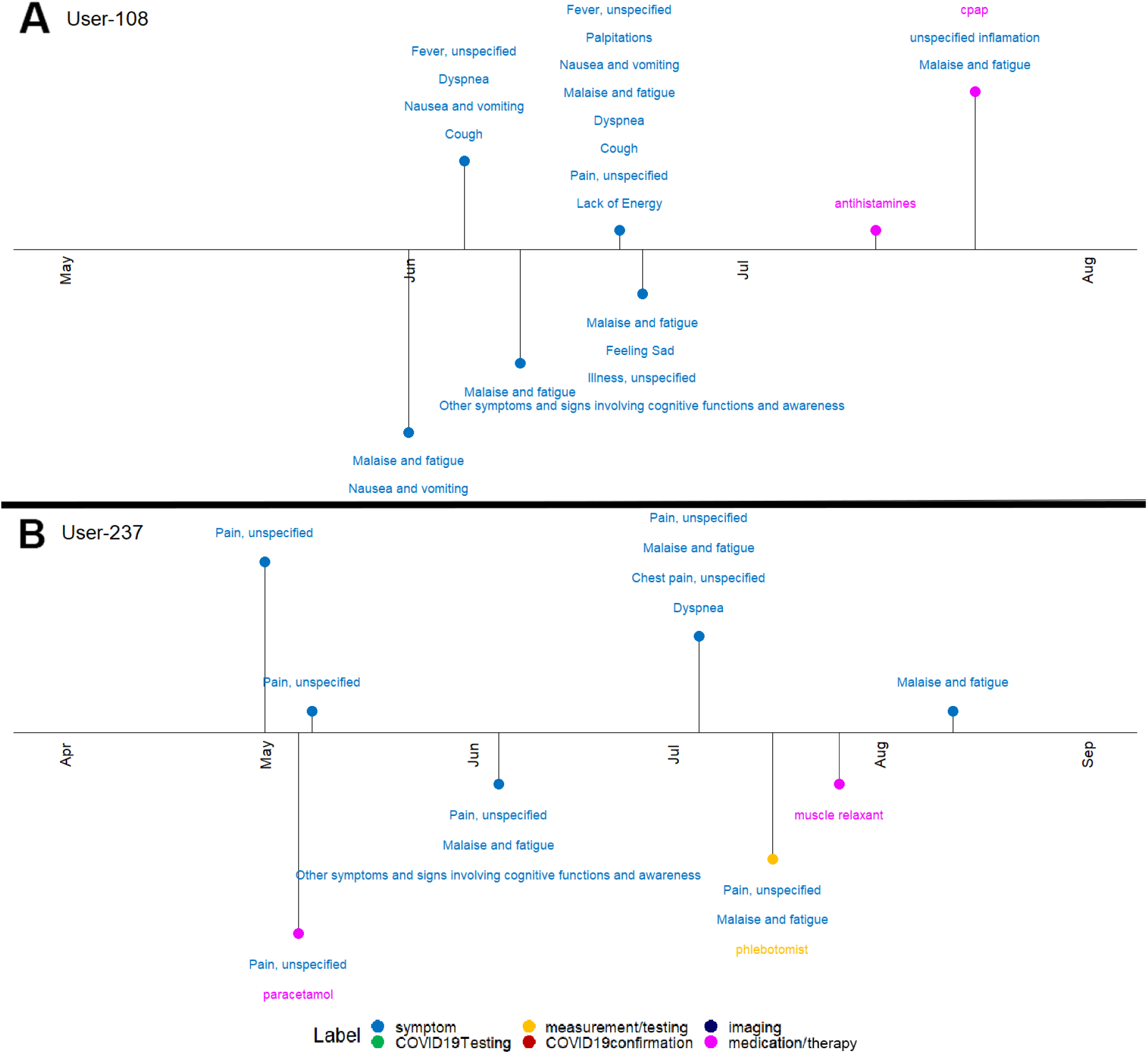
Timeline of annotations of two different long haulers. Note that each dot represents one day of symptoms/medications/testing extracted from their tweets over time.

As we can see from Figure S2, part A and B, all our users’ journeys were very different because symptoms/measurements reported vary, the documentation habits of people and the frequency they posted their experience on Twitter.

Combining all long hauler timelines, we can characterize all users at a granularity of percentage of self-reported symptoms reported on a per-day basis, as seen in Figure S3.

**Figure S3.**
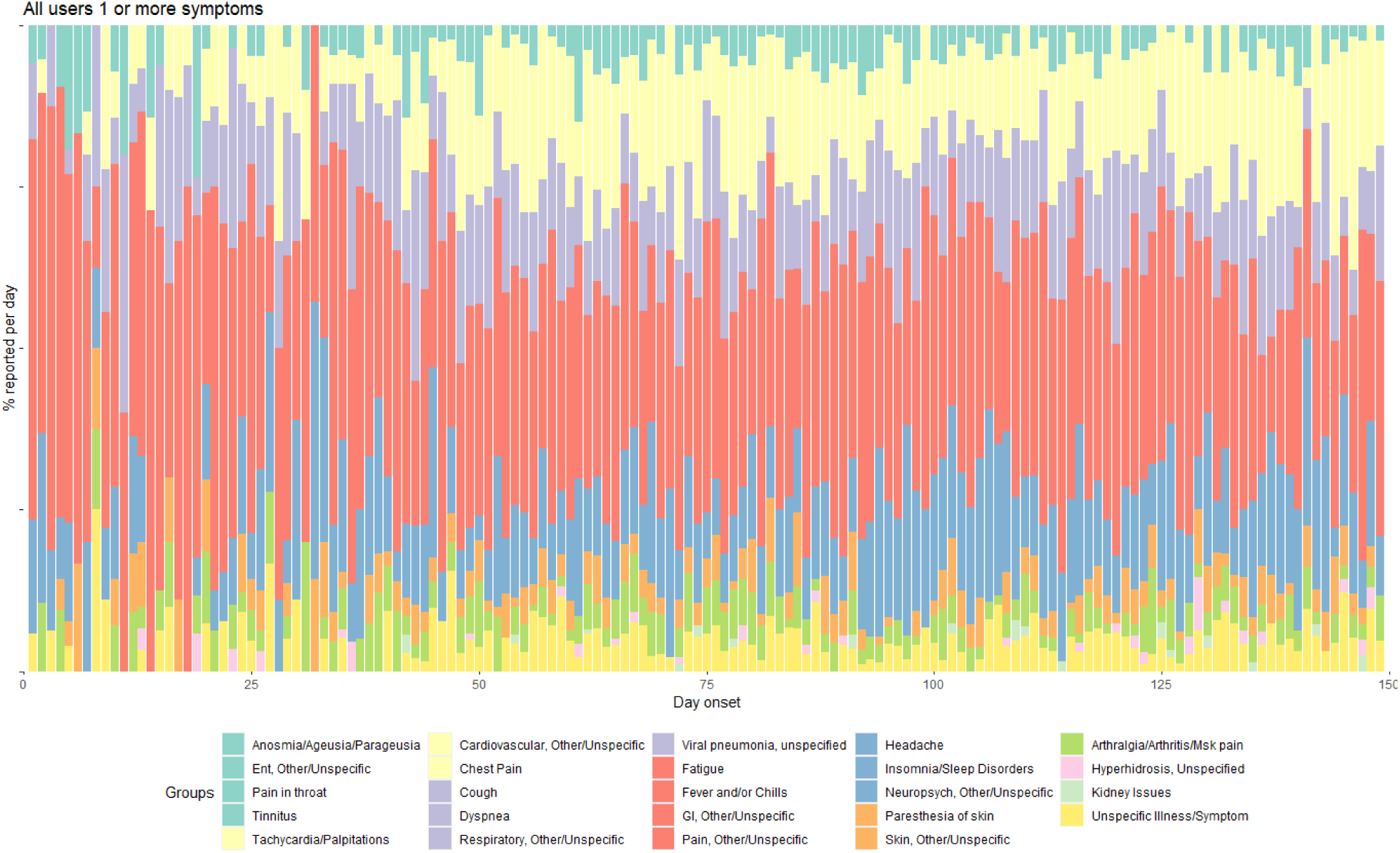
Plot of all symptom groups presented by Twitter users on any day (starting after self-reported diagnosis) during their long-COVID journey. The methods section describes all steps to get to this visualization. GI is used for Gastro-Intestinal and ENT for Ear, Nose and Throat.

### Annotation Normalization

One of the major hurdles of using social media data for any kind of research purpose is that the quality of the language used tends to be lower than in literature or news articles. Since people post messages on the fly or are usually constrained by message sizes (Twitter has a max of 280 characters per message), there is ample use of short-hand and ad hoc abbreviations, as well as the use of colloquialisms or words that are not easily captured by formal controlled vocabularies. In previous studies, we have shown that in the case of finding medication names in COVID-19 chatter ^41^, one would miss around 15% of relevant data if not taken appropriate measures to account for variants in the words used. In order to normalize the annotations made by our clinicians, we used the Observational Health Data Sciences and Informatics (OHDSI) vocabulary that includes a collection of biomedical controlled vocabularies such as: RxNorm, SNOMED-CT, ICD9/10, CPT, among many others. The purpose of doing this is to be able to tie the manual annotators to clinically relevant concepts for downstream analysis. This approach has been shown to be effective when ^41^ dealing with COVID-19 Twitter data. Using Spacy ^42^ and a flattened version of the OHDSI vocabulary that only contains unique strings, we tagged our clinician-generated annotations, resulting in a total of 22,174 concept codes. This is a considerable difference from the number of actual annotated labels, 31,385, mostly due to the previously mentioned word variations, misspellings and abbreviations. Additionally, there are text strings/words that are not in any of the vocabularies from the OHDSI vocabulary version we used. To avoid losing any important data from the annotations, we manually reviewed which annotations did not match to any concept, and when needed we either added a new concept or created a mapping to an already existing concept. Table S3 contains some examples of these new concepts we added, as well as examples of mapped strings to existing terms.

**Table S3.** Annotation Normalization

| Annotated string | concept_id | Type |
| --- | --- | --- |
| brainfog | 66000004 | New Concept |
| covid cheeks | 66000008 | New Concept |
| covid toes | 66000009 | New Concept |
| exhausted | 66000013 | New Concept |
| knackered | 66000013 | Mapped to New Concept - 66000013 |
| body ache | 35843410 | Mapped to MedDRA - General body pain |
| body aches | 35843410 | Mapped to MedDRA - General body pain |
| racing hearts | 36356629 | Mapped to MedDRA - Heart rate high |
| raised hr | 36356629 | Mapped to MedDRA - Heart rate high |

As one can see from Table S3, the majority of terms added are terms that are not found in clinical controlled vocabularies due to their novelty and relatedness to COVID-19 like brain fog, COVID cheeks, and COVID toes. Other terms like ‘exhausted’ and ‘knackered’ need to be added since the current form is not found in the OHDSI vocabulary. The point of adding new terms is to add terms that do not exist or synonyms cannot be found in our vocabulary. For the terms that are synonyms or related terms, like ‘body ache’/’body aches’ and ‘racing hearts’/’raised hr’ are mapped respectively to ‘General body pain’ and ‘Heart rate high’. The manually added term mappings, additional misspellings for certain words, and new concepts total 2,186 new additions to our vocabulary. These are needed to fully convert the manual annotations into standardized concepts. When using the updated dictionary and Spacy, we are left with 40,474 concepts derived from the manual annotations and most importantly, no missing clinician annotation. Lastly, we leveraged internal mapping relationships of the OHDSI vocabulary and other external resources, to map all symptoms/conditions to ICD10 codes when possible for clarity and cohesion of our analysis.

### Organ system groupings

With our analysis focusing on the long haulers and their lingering symptoms/conditions, we wanted to examine the symptoms on two levels. One is the most granulary, and explained above, where we identified in all timelines 136 unique symptoms. To look at these in a higher-level and by organ system we manually curated all these codes into 25 higher-level categories: *Anosmia/Ageusia/Parageusia - Arthralgia/Arthritis/Msk pain - Cardiovascular, Other/Unspecific - Chest Pain - Cough - Dyspnea - Ent, Other/Unspecific - Fatigue - Fever and/or Chills - GI, Other/Unspecific - Headache - Hyperhidrosis, Unspecified - Insomnia/Sleep Disorders – Kidney Issues - Neuropsych, Other/Unspecific - Pain in throat - Pain, Other/Unspecific - Paresthesia of skin - Respiratory, Other/Unspecific - Skin, Other/Unspecific - Tachycardia/Palpitations – Tinnitus - Unspecific Illness/Symptom - Viral pneumonia, unspecified*.

## Data Availability

Data will be made available after formal publication.

## Acknowledgements

JMB was funded by a grant by the National Institute of Aging (3P30AG059307-02S1). DPA is funded through an NIHR Senior Research Fellowship (Grant number SRF-2018-11-ST2-004). VH contribution to this work was carried out with support from National Library of Medicine, National Institutes of Health.

## Author contributions

Conceptualization: JMB, DPA

Data Analysis: JMB, DPA

Data Curation: NA, WA, HA, OA, MA, CA, MC, KF, SG, JJ, LYHL, AL, MAM, EM, DM, AP-U, EGR,GS, VS, AV

Paper Writing: JMB, DPA, VH,AL, LM, MAM, EM, DM, KN, RP, VSP, EGR, GS, VS

## Competing interests

### Data availability

Data will be made available during review upon request and will be made publicly available after formal publication.

### Code availability

Code will be made available during review upon request and will be made publicly available after formal publication

