## Appendix for "Characterization of long-term patient-reported symptoms of COVID-19: an analysis of social media data"

**Appendix: PRISMA flow diagram for long hauler user identification (step 1) and manual tweet annotation (step 2)**

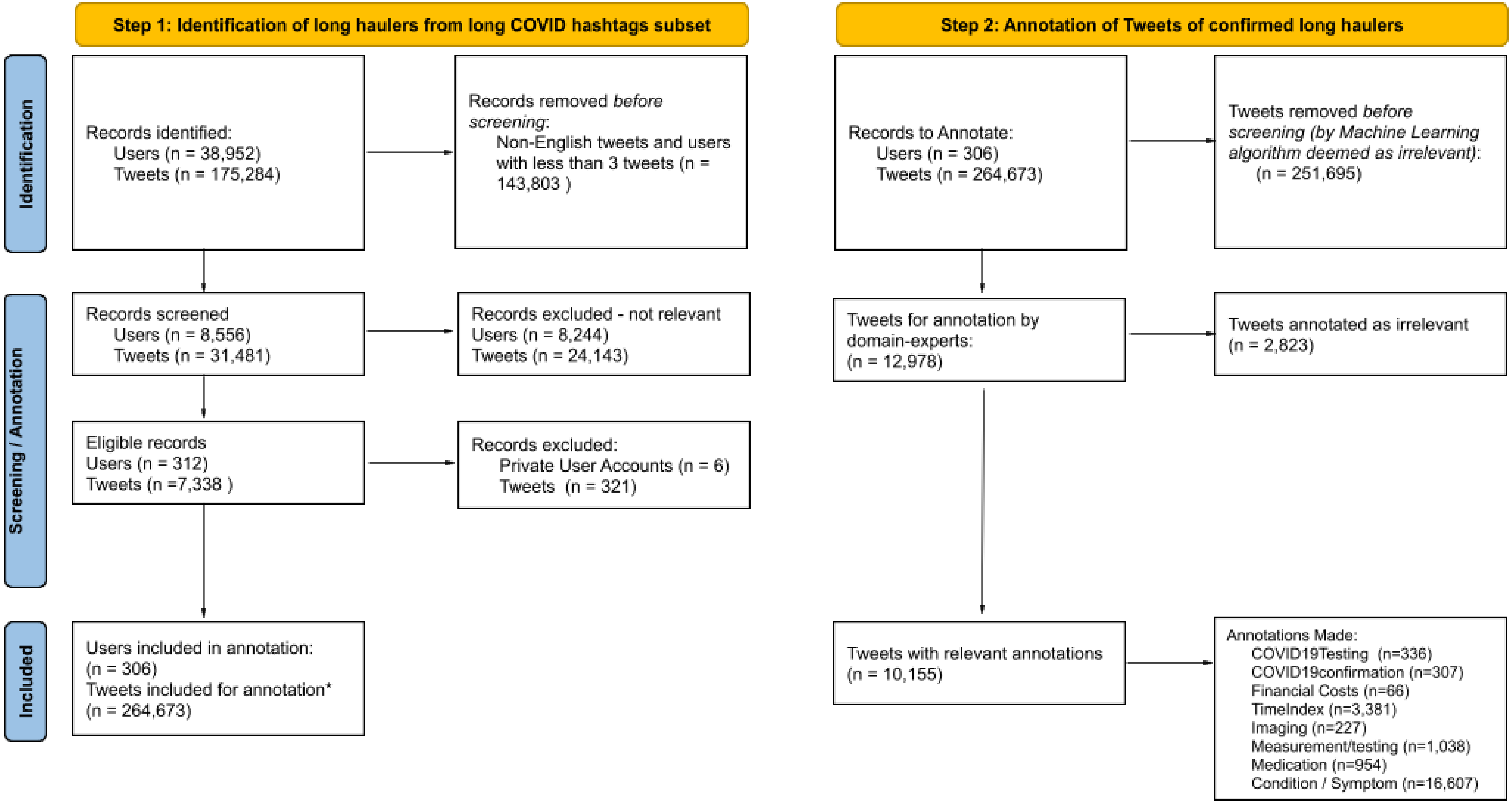


* The number of Tweets here increased from 7,026 to 264,673 as we extracted the complete Twitter timeline tweets (between 2020-01-01 and 2020-08-17) for the 306 users.

*Adapted from:* Page MJ, McKenzie JE, Bossuyt PM, Boutron I, Hoffmann TC, Mulrow CD, et al. The PRISMA 2020 statement: an updated guideline for reporting systematic reviews. BMJ 2021;372:n71. doi: 10.1136/bmj.n71. For more information, visit: [http://www.prisma-statement.org/](http://www.prisma-statement.org/)
